# Efficacy and safety of acupuncture for postpartum hypogalactia: Protocol for a systematic review and meta-analysis

**DOI:** 10.1101/2022.09.10.22279811

**Authors:** Qiong-Nan Bao, Yuan-Fang Zhou, Zi-Han Yin, Qiu-Bi, Hong-Bin Zhao, Zhen-Yong Zhang, Fan-Rong Liang

**Affiliations:** Acupuncture and Tuina School, Chengdu University of Traditional Chinese Medicine, Chengdu, China; Department of traditional Chinese medicine, The First People’s Hospital of Yunnan Province, The Affiliated Hospital of Kunming University of Science and Technology, Kunming, China

## Abstract

**Introduction:** Breast milk is recognised as the best natural food for neonates, but many women experience postpartum hypogalactia (PH). Randomised trials have found that acupuncture exert therapeutic effect on women with PH. However, systematic reviews on the efficacy and safety of acupuncture are still lacking; therefore, this systematic review aims to evaluate the efficacy and safety of acupuncture for PH.

**Methods and analysis:** Eight English databases (PubMed, Cochrane Library, MEDLINE, EMBASE, EBSCO, Scopus, Ovid, Web of Science) and four Chinese databases (China National Knowledge Infrastructure, Wan-Fang, Chinese Biomedical Literature, Chinese Scientific Journal) will be systematically searched from their establishment to 1 September 2022. Randomised controlled trials (RCTs) of the efficacy of acupuncture for PH will be reviewed. The study selection, data extraction, and research quality evaluation will be conducted independently by two reviewers. The primary outcome is the change in serum prolactin level from baseline to the end of treatment. Secondary results include milk secretion volume, total effectiveness rate, degree of mammary fullness, rate of exclusive breastfeeding, infant weight gain, and adverse events. A meta-analysis will be performed using RevMan V.5.4 statistical software. Otherwise, a descriptive analysis will be conducted. The risk of bias will be assessed using the revised Cochrane risk-of-bias tool.

**Ethics and dissemination:** This systematic review protocol does not require ethical approval because it does not include private information/data of the participants. This article will be published in peer-reviewed journals.

**PROSPERO registration number:** CRD42022351849

**Strengths and limitations of this study:**

1. This study will be the first systematic review to evaluate the efficacy and safety of acupuncture for PH by including available clinical trials that have been published in the past.
2. The quality of the included trials will be assessed using the Revised Cochrane Risk of Bias Tool (RoB2) for Randomised Trials.
3. This study will provide more reliable evidence for clinical management because most RCTs on acupuncture for PH have small sample sizes.
4. This study will include only English and Chinese trials, which may lead to missing studies in other languages.

## INTRODUCTION

Postpartum hypogalactia (PH) is a common condition that greatly affects healthy growth and development of babies. It is characterised by little or no breast milk during the lactation period. The World Health Organization recommended that infants should be exclusively breastfed in the first 6 months.^1^ However, only 38% of infants aged under 6 months are exclusively breastfed worldwide, which is lower than the target of 50% in 2025.^2^ In China and Europe, the rate is just 13-39% and 29%, respectively.^3 4^ Many investigators have reported that PH is the most common cause of breastfeeding failure.^5-7^ Breast milk has been identified as the gold standard to provide nutritional support for all infants and prevent infant morbidity and mortality.^8-10^ For mothers, breastfeeding can decrease the risk of breast cancer, ovarian cancer, and type 2 diabetes.^11^ Thus, if a mother’s own milk is insufficient, it poses challenges not only to mothers who desire to provide milk for their babies, but also to infants who need optimal nutrition. Additionally, the cost of not breastfeeding can cause heavy financial burden to households and society; the total global costs of not breastfeeding were estimated at 694322 lives lost annually and economic losses of US$341.3 billion.^12^ Therefore, as the biggest breastfeeding obstacle, PH is a global health priority that needs focused attention and solutions.

Factors contributing to PH include caesarean section, maternal overweight, and poor maternal physical and mental health.^13^ Presently, there are pharmacological and non-pharmacological treatments for PH. The most commonly used pharmacologic are galactagogues such as metoclopramide, domperidone, and sulpiride,^14^ which are not recommended by the Academy of Breastfeeding Medicine,^15^ may increase milk volume with low-certainty evidence,^16^ and are associated with multiple side effects.^17^ Other than drugs, PH can be treated by relaxation techniques, skin-to-skin holding, hand expression, etc.,^18-20^ but the role of these methods in augmenting breast milk might be unsatisfactory. Consequently, it is critical to seek new safe and effective therapies for treating PH.

Acupuncture, a complementary and alternative medicine technique, has been used to treat PH in China for more than 2000 years.^21^ In western countries, acupuncture is one of the most frequently recommended interventions to increase maternal milk production.^22^ Acupuncture involves insertion of fine needles into certain acupoints on the surface of the body, with specific operational methods to restore the balance or harmony of the individual and thereby restore health.^23^ A recent overview^24^ found that acupuncture showed a large effect in increasing lactation success rate post-delivery. Furthermore, several studies have demonstrated that acupuncture is an effective treatment for PH. For instance, Suwikrom et al.^25^ found that acupuncture was superior to conventional treatment for boosting breast milk production in women with PH. Moreover, Su et al.^26^ found that electroacupuncture can markedly improve the breast milk volume and nutrient composition level in women with PH. The mechanisms underlying the effects of acupuncture may involve adjusting microbial and hormonal signals and regulating the activity of the hypothalamic dopamine system and noradrenaline system.^27^ Thus, acupuncture may be a viable option for women with PH.

Many previous randomised controlled trials (RCTs) on acupuncture for PH exist.^25 26 28-30^ However, the clinical outcomes reported vary and the quality of evidence derived from RCTs is unclear, which may lead to inconsistent consensus and risk of bias. Taking these limitations into account, we will conduct a systematic review and meta-analysis of RCTs evaluating acupuncture in PH treatment to solve the above issues and provide more reliable evidence for clinical application and reference for future research.

## METHODS

### Study registration

This study has been registered on PROSPERO (No. CRD42022351849). This protocol is developed and reported according to the Preferred Reporting Items for Systematic reviews and Meta-Analyses Protocols (PRISMA-P) statement.^31^

### Criteria for inclusion of studies in this review

#### Types of studies

All RCTs of acupuncture therapy for PH will be included in the review without any limitations on language and publication time. Non-RCTs, quasi-RCTs, case series, comments, conference abstracts, guides, letters, notes, book sections, editorials, reviews, and animal experimental studies, as well as expert experiences will be excluded.

Types of participants RCTs involving participants with PH, including primipara and multipara participants, will be included without restrictions on age and parity. Studies that include participants with other disorders, such as breast engorgement, mastitis, and breast abscess, will be excluded.

#### Types of interventions

Acupuncture is defined as needle insertion at acupuncture points, Ashi points, or meridian locations and includes manual acupuncture, electroacupuncture, auricular acupuncture, and scalp acupuncture. Studies with interventions that do not use needles to penetrate the skin, such as transcutaneous electrostimulation or acupressure, will be excluded.

#### Types of comparator (s)/control

The following comparative studies will be included:

1. Acupuncture versus placebo acupuncture.
2. Acupuncture versus conventional drugs.
3. Acupuncture with herbs versus herbs alone.
4. Acupuncture in addition to active treatment versus active treatment alone.
5. Acupuncture versus waiting list.
6. Acupuncture versus routine care.

Studies comparing the clinical efficacy between different acupoints, different acupuncture methods, or comparing acupuncture with other complementary and alternative therapies will be excluded.

#### Types of outcome measures

##### Primary outcome

Changes in serum prolactin levels from baseline to the end of treatment will be measured.

##### Secondary outcomes

1. Milk secretion volume
2. Total effectiveness rate
3. Degree of mammary fullness
4. Rate of exclusive breastfeeding
5. Infant weight gain
6. Adverse events

### Search methods for identification of studies

#### Electronic search

The following 12 databases will be searched from their inception to 1 September 2022: PubMed, Cochrane Library, MEDLINE, EMBASE, EBSCO, Scopus, Ovid, Web of Science, China National Knowledge Infrastructure (CNKI), Chinese Biomedical Literature Database (CBM), Chinese Scientific Journal Database (VIP Database), and Wan-Fang Database. The following medical search headings (MeSH) will be used: “acupuncture,” “acupuncture therapy,” “acupuncture points,” “electroacupuncture,” “lactation,” “lactation disorder,” “milk, human,” “milk ejection,” “breast feeding,” “postpartum period,” “randomized controlled trial,” “controlled clinical trial,” and “random allocation.” Chinese databases will be searched using the Chinese translations of these search terms. The search strategy for PubMed is shown in Table 1.

**Table 1.** Search strategy for PubMed

| **Table 1 Search strategy for the PubMed** |  |
| --- | --- |
| **No.** | **Search terms** |
| #1 | Acupuncture. Mesh |
| #2 | Acupuncture. ti, ab |
| #3 | Acupuncture therapy. Mesh |
| #4 | Acupuncture therapy. ti, ab |
| #5 | Acupuncture points. Mesh |
| #6 | Acupuncture points. ti, ab |
| #7 | electroacupuncture. Mesh |
| #8 | electroacupuncture. ti, ab |
| #9 | (body acupuncture or manual acupuncture or auricular acupuncture) . ti, ab |
| #10 | #1 or #2-9 |
| #11 | Lactation. Mesh |
| #12 | Lactation disorder. Mesh |
| #13 | Milk, human. Mesh |
| #14 | Milk ejection. Mesh |
| #15 | Breast feeding. Mesh |
| #16 | Postpartum period. Mesh |
| #17 | (lactation or milk or breast feeding or breastfeeding or hypogalactia or galactosis or colostrum or postpartum or postnatal or puerperal). ti, ab |
| #18 | #11 or #12-17 |
| #19 | randomized controlled trial. pt |
| #20 | controlled clinical trial. pt |
| #21 | random allocation. Mesh |
| #22 | clinical trial. pt |
| #23 | randomized. ti,ab |
| #24 | randomly. ti,ab |
| #25 | trial. ti |
| #26 | #19 or #20-25 |
| #27 | #10 and #18 and #26 |

#### Searching other resources

Potential eligible studies will be manually searched by screening the conference papers and the reference lists of the relevant reviews and the included articles.

### Data collection and analysis

#### Selection of studies

First, all retrieved articles will be imported into EndNote (V.X9). After removing duplicates, two reviewers (Q-NB and Y-FZ) will independently screen the titles and abstracts of all studies to filter out articles that meet the inclusion criteria. Then, the same two reviewers will independently read full text of these articles to determine eligibility; studies that have been excluded will be documented with the reasons for their exclusion. When multiple articles describe the same trial, only the study with the most complete data will be included. Last, the selection results will be cross-checked by the same two reviewers. If there are any disagreements in the process, the final decision will be arbitrated by a third reviewer (Z-HY). The primary selection process is shown in the PRISMA flow chart (Figure 1).

**Figure 1.**
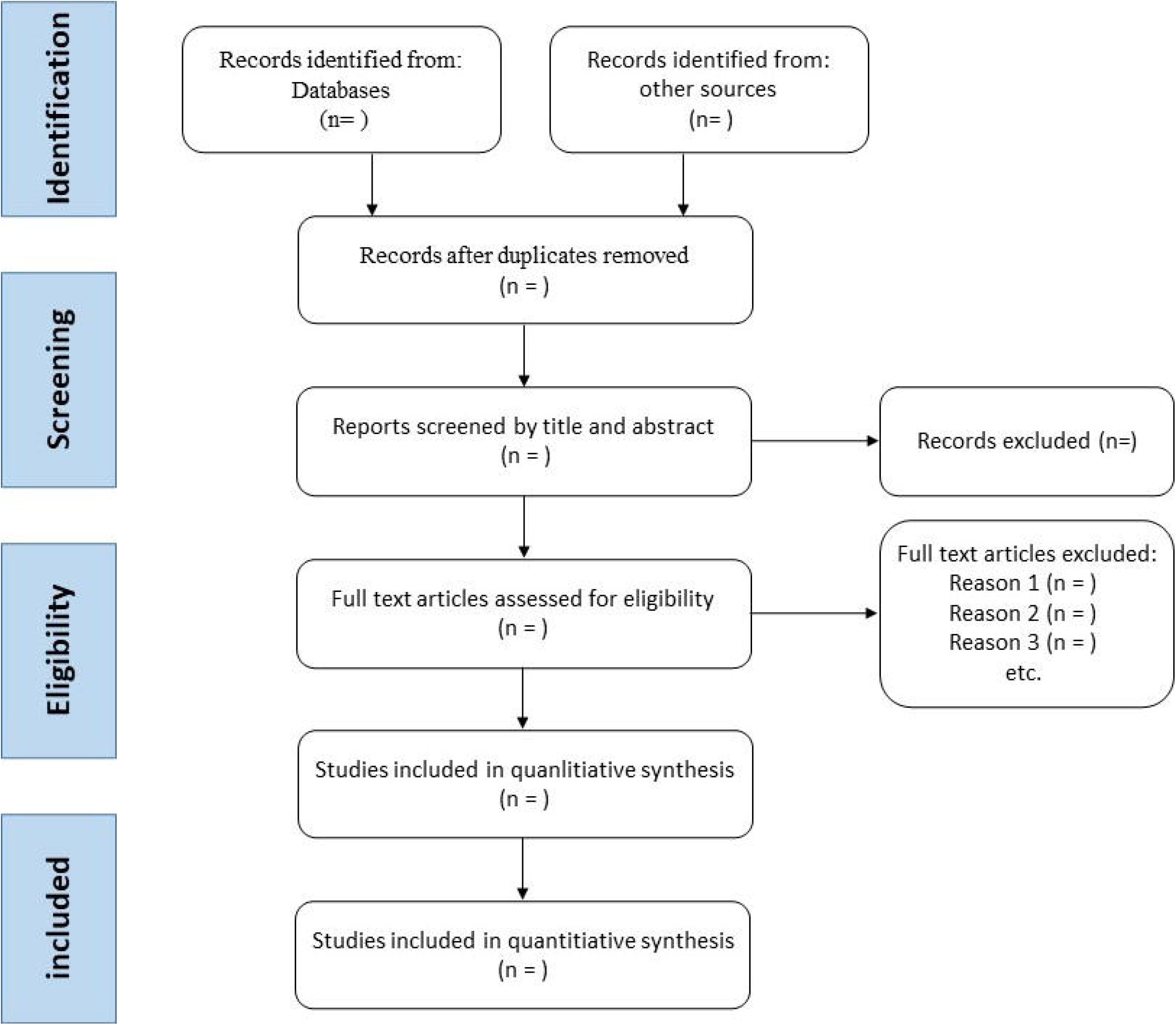
Flow diagram of the study selection process.

#### Data extraction and management

Two reviewers (Q-NB and Y-FZ) will independently extract predefined information from the included studies by using Microsoft Excel. The predefined information will include: publication year, demographic characteristics of the participants, sample size, randomisation, allocation concealment, blinding methods, funding, diagnostic criteria, inclusion/exclusion criteria, details of the intervention and control, outcome measurement methods, follow-up duration, adverse events, and outcomes. If the reported data are missing or unclear, the reviewers will contact the corresponding author to make the supplementation or clarification. Then the same two reviewers will cross-check the accuracy of these data. Any disagreements will be resolved through discussion or judged by a third reviewer (Z-HY).

#### Assessment for risk of bias

Two investigators (Q-NB and Y-FZ) will independently appraise the methodological quality of the included trials using the Revised Cochrane Risk of Bias (RoB 2) tool for randomised trials.^32^ This tool included five domains: randomisation process, deviations from intended interventions, missing outcome data, measurement of the outcome, and selection of the reported results. Each domain corresponds to different signalling questions. According to the signal answer given to different questions, each bias domain will be judged as “low risk”, “high risk”, or “some concerns”, and the overall bias of risk will be given. A third reviewer (Z-HY) will intervene to reach a consensus if inconsistent results are obtained.

#### Measures for the treatment effect

Efficacy data will be synthesised and statistically analysed in RevMan (V.5.4). Dichotomous data will be calculated by the relative risk (RR) with 95% confidence intervals (CIs), and continuous data will be analysed using the weighted mean difference (WMD) or standardised mean difference (SMD) with 95% CIs. The WMD will be used for the same scale or same assessment tools; SMD will be used for different instruments.

#### Assessment of heterogeneity

χ2 tests will be conducted using RevMan software to investigate the statistical heterogeneity, and *P* < 0.10 will be considered significant, in line with the Cochrane Handbook.^33^ The *I*^2^ values will be calculated to quantify the percentage of heterogeneity in the outcome measures. When *I*^2^ values > 30, 50 and 75%, the heterogeneity between studies will be considered moderate, significant or considerable, respectively.

#### Assessment of reporting bias

A Funnel plot^34^ and Egger’s test^35^ will be performed to detect the publication bias if the number of included studies exceed ten.

#### Data synthesis

We will use RevMan software to conduct data synthesis. The use of a fixed-effects or random-effects model will be determined based on the heterogeneity level. If little or no statistical heterogeneity (*I*^2^ ≤ 50% and P ≥ 0.10) is detected among trials, the fixed-effects model will be used for data synthesis. If significant heterogeneity (*I*^2^ > 50% and *P* < 0.10) exists, the random-effects model will be used. If there is considerable heterogeneity (*I*^2^ ≥ 75%) in the trials, meta-analysis will not be performed, and a descriptive qualitative summary will be provided. If necessary, subgroup analysis will be performed with careful consideration of each subgroup. During the meta-analysis process, a third reviewer (Z-HY) may inspect and monitor the procedures irregularly.

#### Subgroup analysis and investigation of heterogeneity

Subgroup analysis and meta-regression will be undertaken to explore potential sources of the heterogeneity in the previous analysis. If the necessary data are available, subgroup analysis will be conducted according to different types of control group and different acupuncture methods (eg, manual acupuncture, electroacupuncture), meta-regression analysis will be carried out based on sample size and treatment duration.

#### Sensitivity analysis

To test the robustness of the meta-analysis results, we will conduct a sensitivity analysis for the primary outcome according to the sample size and the statistical model. The results of the sensitivity analysis will be presented in summary tables and discussed.

#### Grading the quality of evidence

Two reviewers (Q-NB and Y-FZ) will independently evaluate evidence quality using the Grading of Recommendations Assessment, Development, and Evaluation (GRADE) system. The GRADE system evaluates five aspects: limitations in study design, inconsistency, indirectness, imprecision, and publication bias. The quality of evidence will be graded as “high,” “moderate,” “low,” or “very low” in accordance with the GRADE rating standards.

## ETHICS AND DISSEMINATION

This study does not require ethical approval, since no private information of participants will be involved. The results of the present study will be disseminated in peer-reviewed journals or conference presentations. Important protocol amendments will be documented and updated in PROSPERO.

### Patient and public involvement

Patient priorities, experiences, and preferences were not involved in the development of the research question, outcome measures, study design, or the conduct of this review. The results will not be disseminated to participants.

## DISCUSSION

PH is a public health priority that requires efforts to explore more effective treatments. Many RCTs have indicated that acupuncture is effective in promoting breast milk production of women with PH. However, no systematic review in this field has been published. Consequently, it is necessary to conduct a comprehensive review by adding relevant studies into the analysis. The aim of this review is to provide more robust evidence for acupuncture in the treatment of PH and to inform clinical practice and acupuncture research. Inevitably, this study has some limitations. First, diverse treatment courses, acupoints, and methods of milk measurement used in different studies may run a high risk of heterogeneity. Second, only English and Chinese trials will be selected, which may lead to publication bias.

## Data Availability

All data produced in the present study are available upon reasonable request to the authors

## Author Contributions

Q-NB contributed to the concept of the study, and Y-FZ developed the protocol methodology. H-BZ is responsible for planning the statistical analysis. Z-YZ and F-RL will monitor protocol implementation. The original manuscript was completed by Q-NB and was edited by QB. Q-NB and Y-FZ will independently screen potential studies and extract data from the included studies. Q-NB and Y-FZ will assess the risk of bias and perform the data synthesis. Z-HY will arbitrate any conflicts between reviewers and perform quality checks at all stages of review. All authors have read, provided feedback, and approved the final manuscript.

## Patient consent for publication

Not required.

## Provenance and peer review

Not commissioned; externally peer reviewed.

